# The Impact of Psychology Interventions on Changing Mental Health Status and Sleep Quality in University Students during the COVID-19 Pandemic

**DOI:** 10.1101/2020.09.01.20186411

**Authors:** Jing Xiao, Yu Jiang, Yu Zhang, Xinyi Gu, Wenjing Ma, Bo Zhuang, Ziqi Zhou, Lingli Sang, Yitian Luo, Yulong Lian

## Abstract

**Objective:** We evaluated the change in mental health and sleep quality of college students at four time periods.

**Methods:** Mental health status and sleep quality were using the Pittsburgh Sleep Quality Index (PSQI) and Symptom Checklist-90-Revised (SCL-90-R) questionnaire across four time periods. Psychology interventions were carried out from the third period.

**Results:** Students in the third period had higher PSQI total scores [mean (SD), 6.01 (3.27)] than those in the first period [5.60 (3.11)], second period [4.17 (2.10)] and fourth period [4.09 (2.80)]. After adjustment for covariates there was a decline of 1.89 points in the PSQI in the fourth period compared with the highest period. The SCL-90-R scores were highest in the second period [121.19 (47.83)], and were higher than the scores in the first [107.60 (52.21)] and second period [107.79 (27.20)] and lowest in the fourth period [97.82 (17.12)]. The decline in scores was 23.38 points after adjustment for covariates. The prevalence of psychological distress and sleep disturbances respectively decreased from 28.6% to 11.7% and from 10.4% to 2.6% comparing to the highest period. Sleep quality showed a significant positive correlation with mental health status.

**Conclusions:** The pattern of change in mental health status was different to that of sleep quality. The implementation of comprehensive psychology intervention may improve mental health and sleep quality. These findings may inform public health policy during the reopening of schools in other regions.

## Introduction

The COVID-19 pandemic has resulted in more than 15300 thousand cases and 630 thousand deaths to date worldwide(1). The ongoing COVID-19 pandemic not only has the risk of death due to the viral infection but is also an unprecedented unique event in terms of the physical and mental health of the general public (2). In response to COVID-19, countries around the world have closed schools. As the number of COVID-19 cases and related deaths decline, countries are now considering reopening schools. Given that most new COVID-19 cases in China are imported, the country gradually started to reopen schools from mid-April. A series of multifaceted public health interventions have been suggested for schools in China to limit the spread of COVID-19. The COVID-19 pandemic, and the actions taken in response to the pandemic in schools, will have far-reaching consequences on mental health and sleep quality in students, A previous study in China indicated that 24.9% of college students experienced anxiety due to the COVID-19 outbreak. Of these students, 0.9% experienced severe anxiety, and 21.3% experienced mild anxiety (3). Another study showed that the stay-at-home order contributed to reduced social activity and changed sleep behavior in 139 university students during the COVID-19 pandemic (4). Sara M. reported later bed time, sleep latency, wake-up time and depressive and anxiety symptoms during the COVID-19 emergency (5). Several studies have also reported the psychological consequences of strict quarantine measures (6). In addition, these interventions inevitably have a complex effect on sleep health.

According to the trends in the COVID-19 pandemic and the reopening of schools in China, some psychology measures were also proposed by the National Health Commission of China (7) and the Ministry of Education of the People’s Republic of China (8). These psychology interventions for college and university students included, but were not limited to psychological status assessment, social support, on-line and on-site psychological counseling, and psychological health education. These measures were expected to change the mental health and sleep health of college and university students. However, to our knowledge, no study has yet comprehensively evaluated the changes in mental health and sleep health in university students during different periods of the COVID-19 pandemic, and the impact of these psychology interventions on mental health and sleep quality.

In this study, we investigated and analyzed the change in mental health status and sleep quality of university students during the COVID-19 pandemic according to key events and psychology interventions, and the effect of these psychology interventions. Understanding such changes and the impact of psychology interventions could help inform public health recommendations with the goal of improving physical and mental health during the COVID-19 pandemic.

## Methods

### Study population

We conducted an observational study to analysis changes in multiple dimensions of sleep quality and mental health during different time periods of the COVID-19 pandemic. Three classes were sampled using cluster sampling methods in Nantong University, Jiangsu province, China. All 92 students were invited to participate in the study. Eighty-six students responded to the questionnaire, and 9 students dropped out of the study. Sleep quality and mental health data were collected four times from January 27 to July 3, 2020 in the same university students. Seventy-seven students were included in the final analysis. The research protocol was approved by the Ethics Committee of Nantong University. Written informed consent was obtained from all participants.

### Key reopening school events and public health interventions in the four periods

To better reflect the change in sleep quality and mental health, and the corresponding psychological intervention effects, four time periods were studied based on dates of the assessment of participants, change in public health emergency level and key events in the reopening of schools. 1) First period (January 27 to February 28, 2020): Public health response level I was activated in January 27, 2020 in Jiangsu province, China. During this period, the government complied with strict public health interventions (mainly including treatment and isolation of COVID-19 patients, traffic restriction, social distancing, universal symptom survey by community workers and volunteers, closure of all public places, home quarantine and centralized quarantine for presumed cases, those with respiratory symptoms and close contacts, wearing of face masks in public places and personal hygiene such as hand washing and disinfection). Students were at home for the winter holiday. The new semester started on February 24 and students began on-line classes at home. There were no new confirmed domestic cases from February 19. From February 25, 2020, Jiangsu province downgraded their public health emergency response level (Level II) (mainly including opening traffic restrictions, tighten up border quarantine, nucleic acid testing on inbound arrivals, resumption of work, social distancing, reopening of public places, voluntary wearing of face masks in public places and personal hygiene such as hand washing and disinfection). The participants were assessed on February 28, 2020 (the first time). 2) Second period (March 28 to April 27, 2020): From March 28, 2020, Jiangsu province downgraded the public health emergency response level(Level III), and focused on preventing inbound cases and domestic resurgence (mainly including tightening-up border quarantine, nucleic acid testing on inbound arrivals, fully restoring normal order in work and daily life). Students returned to school on April 17, 2020 and were educated by a mixture of on-line and in-person classes. School-related measures were implemented including staying at school, wearing masks, hygiene and environmental cleaning, screening and management of sick students, daily self-monitoring of body temperature and symptoms and physical distancing. Also comprehensive mental health interventions were carried out including psychological status assessment, social support, on-line and on-site psychological counseling and psychological health education. The second assessment of mental health and sleep quality was on April 27. 3) Third period (April 28 to May 28, 2020): Jiangsu province maintained public health emergency response level III and public health measures and school-related measures were the same as those during the second period. The third assessment of students was on June 2. 4) Fourth period (June 3 to July 3, 2020): the government maintained public health emergency response level III. School-related measures were the same as those during the third period. In this period, students finished their final examinations ready for the summer vacation. The fourth assessment was on July 3 (Figure 1).

**Figure 1.**
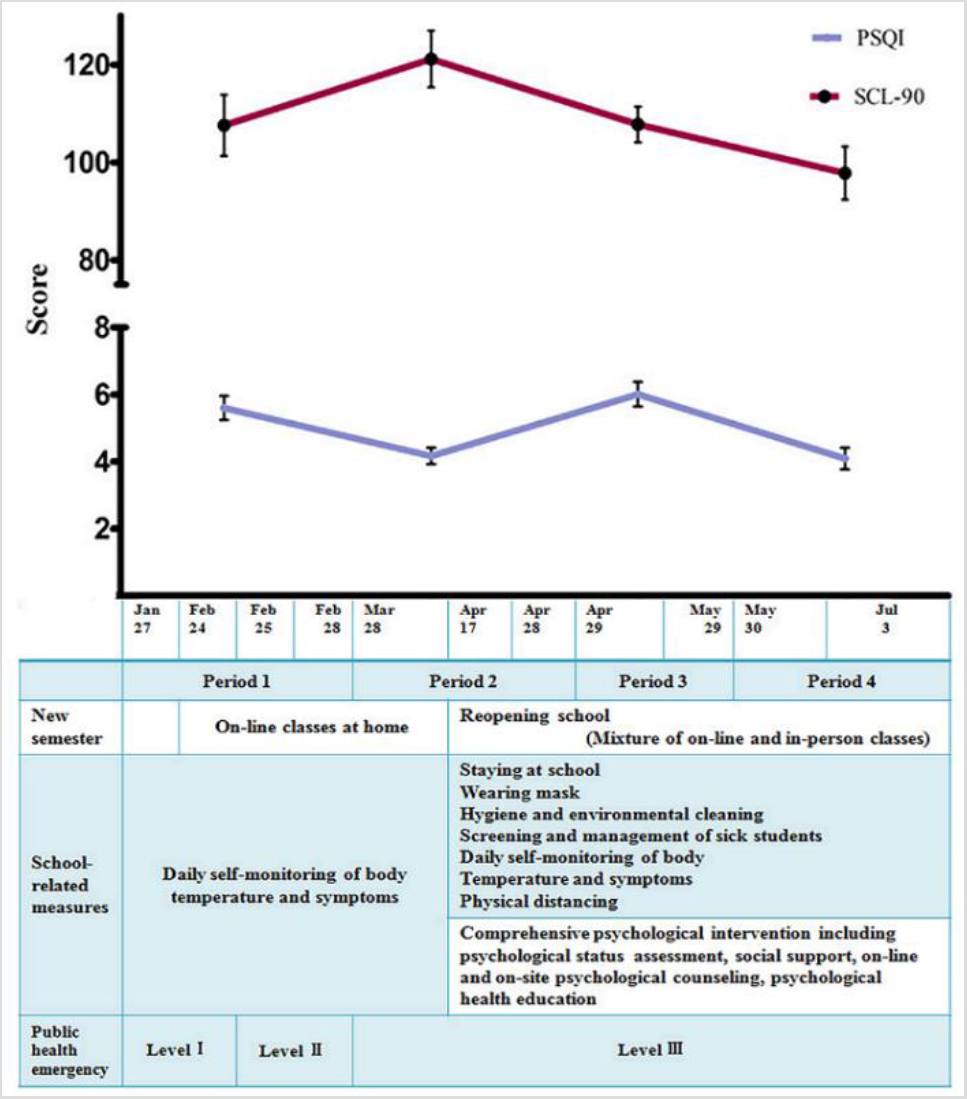
The changing mental health and sleep quality of students, key school-related events and public health emergency level across the 4 periods during the COVID-19 pandemic in Jiangsu province China

Comprehensive psychology interventions were implemented by the school according to the guidelines on COVID-19 prevention and control in higher education institutes proposed by the Ministry of Education of the People’s Republic of China (8). These included psychological status assessment following return to school, social support from the teacher, instructor and students’ association union in life and emotional events. Psychological counseling was included on-line (such as WeChat, Tencent QQ, and 24 h psychological crisis intervention hotline) and on-site counseling was carried out by a psychology consultation teacher. Psychological health education was implemented regularly by the school using on-line classes and internet apps regarding knowledge of COVID-19, adjustment skills related to trauma and stress, and relaxation therapy, etc.

### Sleep quality

The PSQI is a 19-item self-rated questionnaire designed to measure sleep quality during the previous month (9). The PSQI consists of multiple dimensions of sleep quality including subjective sleep quality, sleep latency, sleep duration, sleep efficiency, sleep disturbance, use of sleep medication, and daytime dysfunction. Each domain ranges from 0 to 3. Individual scores were summed to obtain a global score ranging from 0 to 21, with higher global scores indicating poor sleep quality. A PSQI global score ≥8 was defined as sleep disturbance in Chinese undergraduate students (10,11). The Chinese version of the PSQI has shown good validity and reliability (10).

### Mental health status

We measured the general mental health of participants using the SCL-90-R which is a self-administered questionnaire used to measure mental health status and psychological symptoms, and this questionnaire has been used in several community-based epidemiology studies (12). It consists of 90 items across nine primary symptom domains: (a) somatization, (b) obsessive-compulsive, (c) interpersonal sensitivity, (d) depression, (e) anxiety, (f) hostility, (g) phobic anxiety, (h) paranoid ideation, and (i) psychoticism. Participant responses were measured using a five-point Likert-type scale ranging from 0 (“asymptomatic”) to 4 (“very severe”). The Chinese version of the SCL-90-R has high reliability and validity (13), and psychological distress was defined as a global severity index score corresponding to a T-score at least 63 (12).

### Covariates

There were several confounding variables, including gender, age and family address. Body weight and height were measured to calculate body mass index (BMI in kg/m^2^). Information regarding cigarette smoking, alcohol consumption and physical exercise was also collected. Having isolated neighbors and cases in the community were also asked. Chronic disease was assessed by the question ‘‘do you have medically-confirmed chronic diseases?’’. The answer was “no” or “yes”.

### Statistical analysis

Statistical analyses were performed using SAS Version 9.1 for Windows. An analysis of descriptive statistics was conducted to illustrate the demographics and other selected characteristics of the participants. We used a mixed linear model that has been recommended for repeated measures to compare the changes in sleep quality and mental health at different time periods. Considering the bi-directional relationship between mental health and sleep quality previously reported (14), we used a mixed linear model which can analyze correlations between two dependent variables proposed by Roy A (15), to analyze the relationship between sleep quality and mental health in different periods, and the correlation between mental health and sleep quality including covariates (gender, BMI, physical exercise, chronic disease, having isolated neighbors or COVID-19 cases in the community, family address). As none of the participants had a smoking or drinking behavior, these covariates were not used for analysis.

## Results

The demographics of the participants are shown in Table 1. The participants consisted of 62 (80.5%) women and 15 (19.5%) men. The mean age was 20.97 (SD, 1.24) years. Most participants(90.9%) did not have isolated neighbors or cases who were infected with COVID-19 in the community.

**Table 1.**
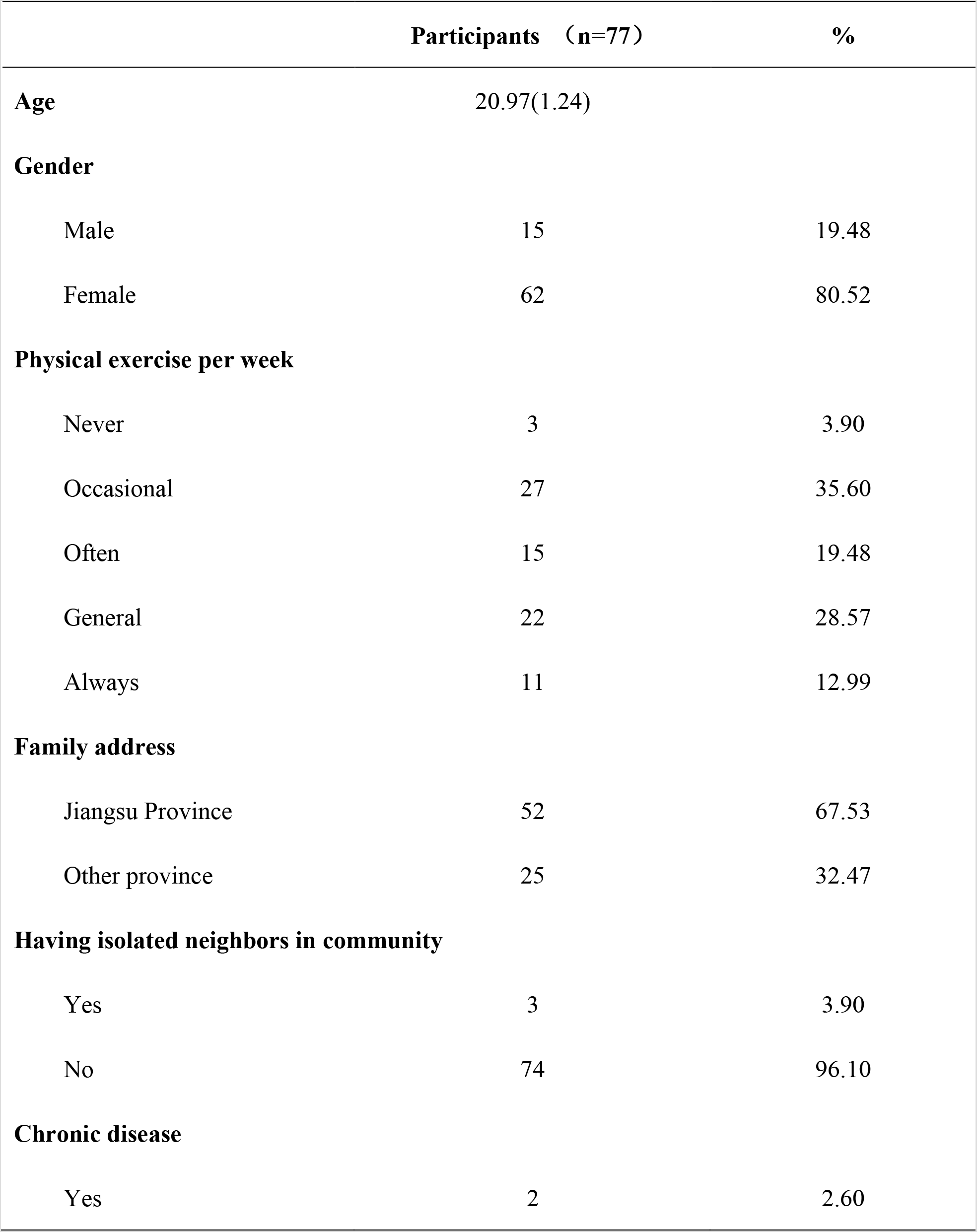
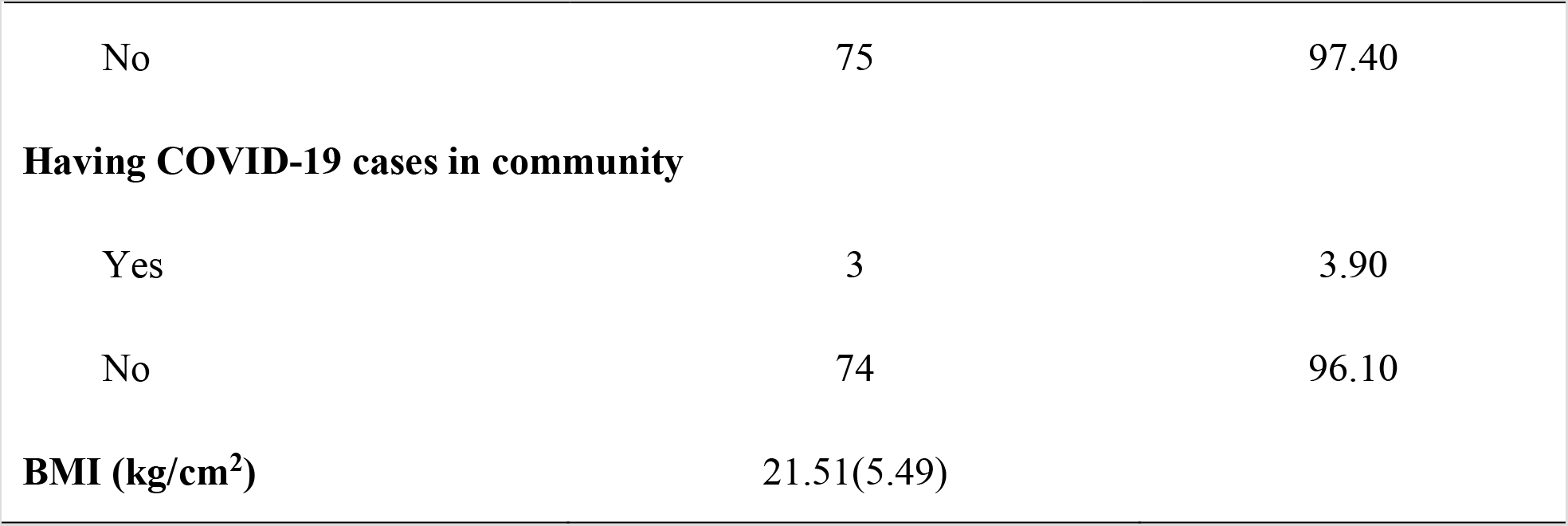
Sociodemographic characteristic of participants

A mixed linear model was used to compare the changes in SCL-90-R and PSQI scores in the different time periods (Table 2). We found significant differences in the overall scores of the PSQI and SCL-90 in these four periods. PSQI total scores in the third period were significantly higher [mean (SD), 6.01 (3.27)] than those in the second [4.17 (2.10)] and fourth period [4.09 (2.80)]. Subjects in the first period had significantly higher PSQI scores [mean (SD), 5.60 (3.11)] than those in the second and fourth period. In the first period, subjective sleep quality and sleep latency showed higher scores. In the second period, the score for sleep duration was higher. Daytime disturbance was higher in the third period. The SCL-90-R scores in the third period were significantly higher [121.19 (47.83)] than those in the first [107.60 (52.21)], second [107.79 (27.20)] and fourth period [97.82 (17.12)]. Subjects in the third period had significantly higher SCL-90-R scores than in the fourth period. Subjects in the second period had the highest scores in the following symptoms: somatization, obsessive-compulsive, interpersonal sensitivity, depression, hostility, phobic anxiety, paranoid ideation and psychoticism (P< 0.05). However, the mental health symptoms decreased in the fourth period. The prevalence of sleep disturbance and psychological distress was 26.0% and 6.5% in the first period, 6.5% and 10.4% in the second period, 28.6% and 6.5% in the third period, and 11.7% and 2.6% in the fourth period.

**Table 2.**
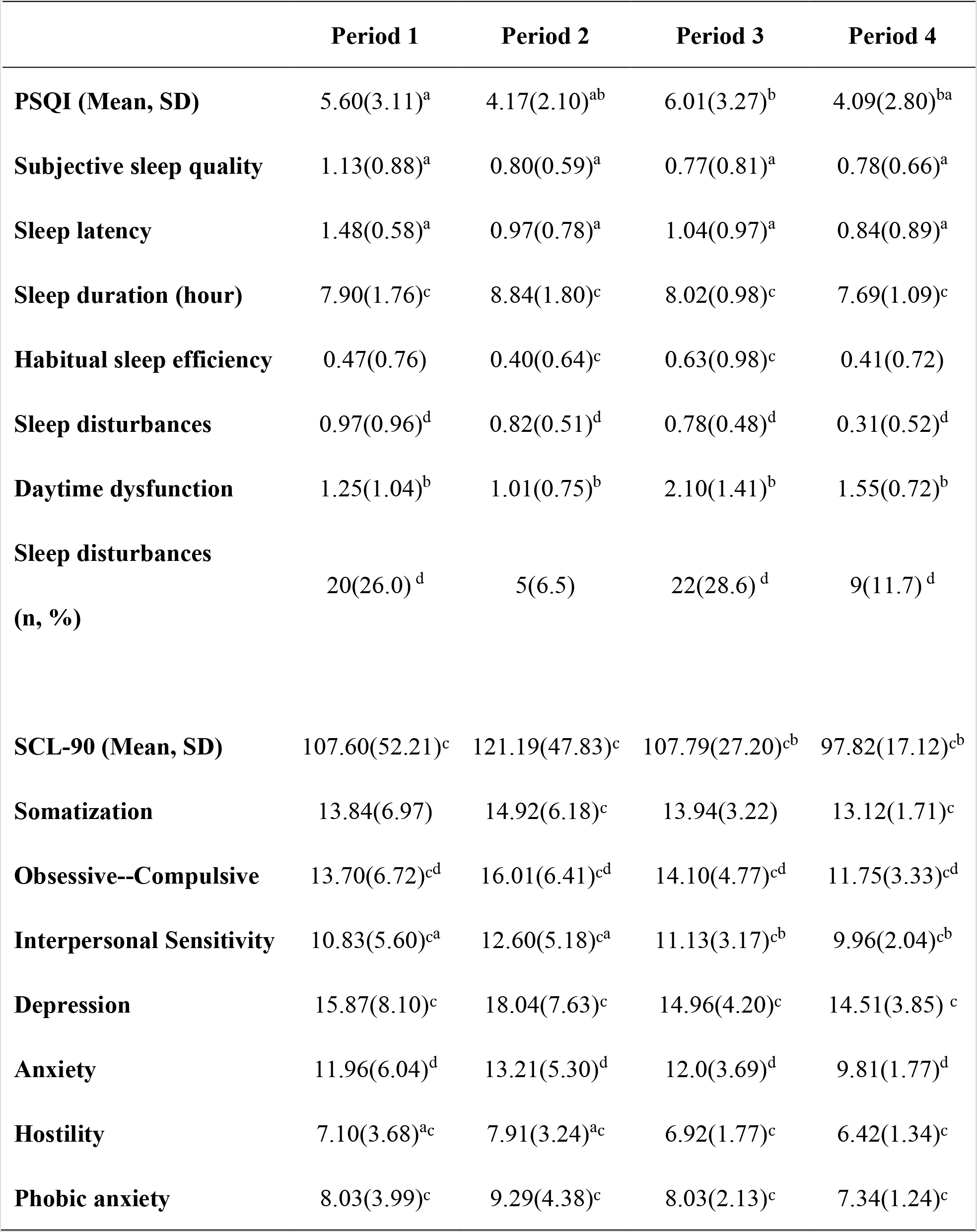
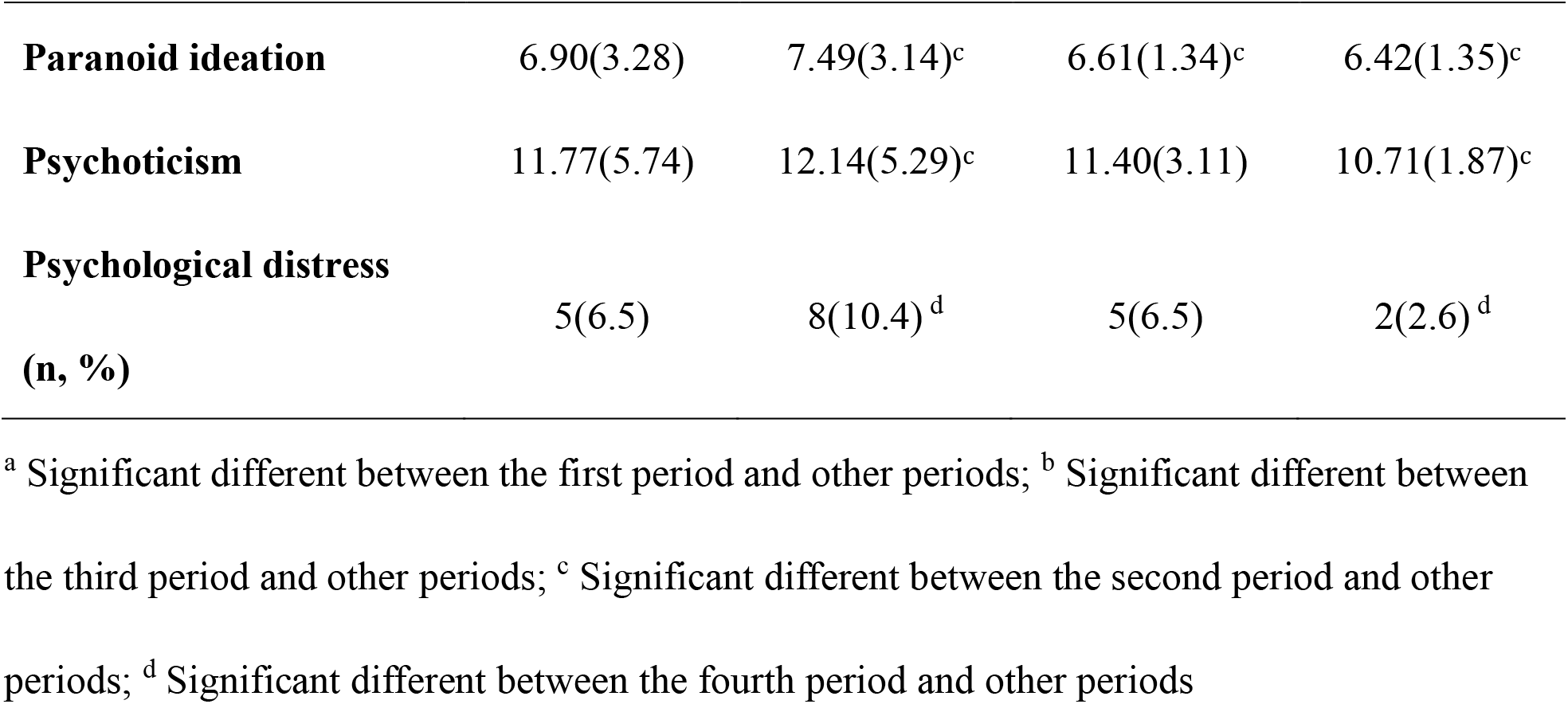
Comparison of SCL-90-R score and sleep quality in the four periods

There was a positive association between the SCL-90-R scores and PSQI [ρ = 0.33, 95%CI (0.12–0.57)]. Participants in the second and fourth period had PSQI scores 1.86 and 1.89 points lower than those in the third period (highest score), respectively. Differences in PSQI scores were not found between the third and first period. Participants in the second period had SCL-90-R scores 13.13, 13.10 and 23.38 points higher than those in the first, third and fourth period, respectively (Table 3 and Figure 2).

**Table 3.** Estimated fixed effects of periods on mental health and sleep quality adjusted for covariates.

|  | Sleep quality |  |  |  | SCI-90-R |  |  |  |
| --- | --- | --- | --- | --- | --- | --- | --- | --- |
| | $\beta$ | SE | 95%CI | | $\beta$ | SE | 95%CI | |
| Intercept | 5.83<sup>a</sup> | 0.36 | 5.12 | 6.54 | 107.90<sup>a</sup> | 6.34 | 95.26 | 120.54 |
| Period 1 | -0.15 | 0.54 | -1.24 | 0.93 | -13.13<sup>a</sup> | 4.81 | -22.71 | -3.54 |
| Period 2 | -1.86<sup>a</sup> | 0.44 | -2.73 | -0.99 | 0 |  |  |  |
| Period 3 | 0 |  |  |  | -13.10<sup>a</sup> | 6.56 | -26.17 | -0.03 |
| Period 4 | -1.89<sup>a</sup> | 0.50 | -2.89 | -0.89 | -23.38<sup>a</sup> | 3.90 | -35.32 | -11.43 |
| Correlation coefficient (PSQI; SCL-90) $\rho=0.33^a$ 95%CI (0.12-0.57) | | | | | | | | |

<sup>a</sup> P<0.05 Adjusting for gender, BMI, physical exercise, chronic disease, having isolated neighbors or COVID cases in the community, and family address.

**Figure 2.**
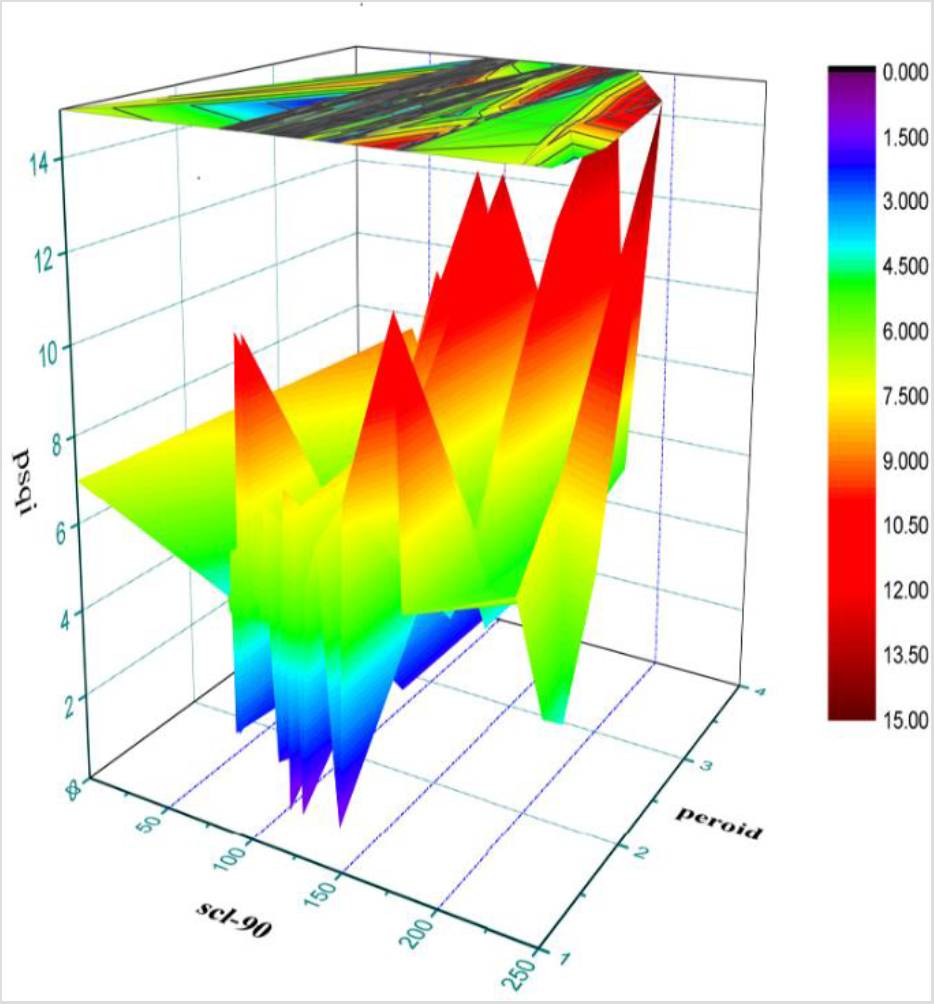
The relationship between period, mental health and sleep quality

## Discussion

To our knowledge, this is the first study to assess the mental health and sleep quality of university students during different time periods of the COVID-19 pandemic. Mental health had a positive association with sleep quality, and comprehensive psychology interventions can improve the mental health status and sleep quality of students.

Several previous studies have suggested that public health emergencies may significantly affect the psychological health of college students, and can increase anxiety, fear, and worry (3–5). We found that students showed the highest mental health symptoms and the prevalence of psychological distress three months after the COVID-19 outbreak, which is in agreement with the findings reported during the previous SARS-CoV-1 epidemic where the general public had psychiatric symptoms months after the epidemic was controlled (16). Participants also showed greater symptoms, such as somatization, obsessive-compulsive, interpersonal sensitivity, depression, hostility, phobic anxiety, paranoid ideation and psychotic symptoms. Although the local public health emergency level declined from I to III and no Covid-19 cases were identified in the local province, the schools announced that there would be no in-person, on-campus option to attend classes, and students were to complete on-line courses at home ready to return to school and adapt to the school measures after the initial stage of school reopening. These measures and events may have led to more stress in students. One recent study has also indicated that students were more sedentary, with increased phone usage and reported increased mental health symptoms related to prolonged isolation time and subsequent academic breaks (17). After the psychology interventions, mental health status and related symptoms immediately declined and decreased to the lowest levels after two months. This indicated that psychology interventions had a beneficial effect on poor mental health within a short time during the COVID-19 pandemic. Considering that approximately 70% of individuals with mental disorders are aged less than 25 years (18), effective psychology interventions should be applied in these students for life-long benefits during the pandemic. Also effective long-time surveillance of mental health and addressing the early onset of mental disorders in schools must be an essential component of the school-focused mental health policy during the pandemic.

Students’ sleep quality was affected by the COVID-19 pandemic and improved by the psychology interventions. During the first period, local government complied with the strict public health interventions, such as the compulsory stay-at-home policy for all residents. This COVID-19 ‘lockdown’ may have altered subsequent sleep behavior. Furthermore, the high use of on-line media (particularly social media) among university students also had an additional negative impact on sleep quality during the COVID-19 pandemic (19). Self-reported sleep quality improved noticeably in the second period, which corresponded to widespread policy changes in the local government public health emergency response (from level I to III). However, participants showed poorest sleep quality in the third period, especially habitual sleep efficiency and daytime dysfunction. Of note, during the third period, schools had reopened and students had returned to school. During this stage, school-related interventions were mainly carried out, including stay-at school, mixture of on-line and in-person classes, wearing masks, daily self-monitoring of body temperature, psychological health assessment and education, etc. Therefore, these measures may have resulted in worsening of sleep quality. These poor sleep behaviors may contribute to and worsen major health problems, including cardiovascular disease, obesity, diabetes, mood disorders, and immune disorders. However, following the mental health interventions, we did not find that the interventions had an immediate effect, and two months after the interventions the PSQI declined to the lowest level. This suggested that psychology interventions for sleep may require more time or more sleep hygiene measures.

Sleep quality and mental health conditions not only share common causes but also show a bidirectional relationship. Our findings also agree with a previous report on the bi-directional complex relationship between the prevalence of sleep status and psychological outcomes among healthcare workers during the COVID-19 outbreak (14). Longitudinal studies also show that sleep difficulties before a trauma and in the weeks and months after a trauma are a predictor of the development of posttraumatic stress disorder (20, 21). It is also highly likely that the presence of mental health symptoms leads to worsening of sleep problems. This knowledge including our findings could be used to improve mental health services during the COVID-19 pandemic. It is necessary to integrate sleep interventions into mental health interventions. In the present study, we conducted a range of psychology interventions in the last two periods, such as psychological status assessment, social support, on-line and on-site psychological counseling and psychological health education. We found that students’ sleep quality and mental state markedly improved in the last period. Therefore, it is likely that mental health interventions targeted towards specific populations might promote mental health and sleep quality. Understanding the changes in mental health and sleep quality during the pandemic and the effect of psychology interventions could help inform public health recommendations for preventing the onset of stress-related disorders such as post-traumatic stress disorder in specific groups during the COVID-19 pandemic.

The major strength of this study is the assessment of mental health status and sleep quality using intensive repeated measures. This reliable research design showed detailed changes in dependent variables with time. In addition, the impact of psychology interventions on mental health and sleep quality were analyzed. However, the limitations of the current study should also be considered. First, small sample size may have led to bias our findings. Second, the study was restricted to university students, so the results may not apply to other populations. Third, unfortunately we did not collect data in March, the period of activated public health response level II; thus, we do not know the changes in mental health and sleep status during this time. Finally, some important covariates such as resilience and psychological adjustment were not included in the study, and the improved outcomes may be partly due to these factors.

In conclusion, we found that the pattern of change in mental health status was different to that of sleep quality, and the implementation of comprehensive mental health interventions may improve mental health and sleep quality during the COVID-19 pandemic. We recommend continuous surveillance of mental health and sleep quality, and the early provision of mental health and sleep hygiene interventions for university students when considering reopening schools.

## Data Availability

The data of this study is not available as the data also forms part of an ongoing research.

## Statement of Ethics

All participants provided their online informed consent. The study was approved by the Ethics Committee of Nantong University.

## Acknowledgements

This work was supported by the Natural Science Foundation of Jiangsu Province, China(Grant Number: BK20171256); Qinglan Project of Jiangsu Province of China.

## Notes

### Competing Interest Statement

The authors have declared no competing interest.

### Funding Statement

This work was supported by the Natural Science Foundation of Jiangsu Province, China (Grant Number: BK20171256) and Qinglan Project of Jiangsu Province of China.

### Author Declarations

The study was approved by the Ethics Committee of Nantong University.

## References

1 World Health Organization [Internet]. Coronavirus disease (COVID-19) pandemic;2020.

2 Ghebreyesus TA. Addressing mental health needs: an integral part of COVID-19 response. World Psychiatry 2020;19:129–30.

3 Cao W, Fang Z, Hou G, Han M, Xu X, Dong J, Zheng J. The psychological impact of the COVID-19 epidemic on college students in China. Psychiatry Res 2020;287:112934.

4 Wright KP Jr, Linton SK, Withrow D, Casiraghi L, Lanza SM, Iglesia HD, Celine V, Depner CM. Sleep in university students prior to and during COVID-19 Stay-at-Home orders. Curr Biol 2020;30:R797–R798.

5 Marelli S, Castelnuovo A, Somma A, Castronovo V, Mombelli S, Bottoni D, Leitner C, Fossati A, Ferini-Strambi L. Impact of COVID-19 lockdown on sleep quality in university students and administration staff. J Neurol 2020;1–8. doi: 10.1007/s00415-020-10056-6.

6 Vindegaard N, Benros ME. COVID-19 Pandemic and Mental Health Consequences: Systematic Review of the Current Evidence. Brain Behav Immun 2020;S0889-1591(20)30954-5.

7 Xiang C, Zhibing H, eds. Guidelines on COVID-19 prevention and control in higher school institutes. Ministry of Education of the People’s Republic of China. Beijing: Chinese People’s Medical Publishing House Co., LTD; 2020.

8 National Health Commission of the People’s Republic of China [Internet]. Guidelines on COVID-19 prevention and control in university and college; 2020.

9 Buysse DJ, Reynolds CF 3^rd^, Monk TH, Kupfer DJ. The Pittsburgh Sleep Quality Index: a new instrument for psychiatric practice and research. Psychiatry Res 1989;28:193–213.

10 Guo S, Sun W, Liu C, Wu S. Structural Validity of the Pittsburgh Sleep Quality Index in Chinese Undergraduate Students. Front Psychol 2016;7:1126.

11 Shi SP, Xiong DY, Yan QR. Sleep quality among college students and associated factors. Chin J Sch Health 2013;34:1462–4. Chinese

12 Derogatis LR. SCL-90-R Administration, Scoring & Procedures Manual-II. Towson, MD: Clinical Psychometric Research 1983:14–5.

13 Yu Y, Wan C, Huebner ES, Zhao X, Zeng W, Shang L. Psychometric properties of the symptom check list 90 (SCL-90) for Chinese undergraduate students. J Ment Health 2019;28:213–9.

14 Freeman D, Sheaves B, Waite F, Harvey AG, Harrison PJ. Sleep disturbance and psychiatric disorders. Lancet Psychiatry 2020;7:628–37.

15 Roy A. Estimating correlation coefficient between two variables with repeated observations using mixed effects model. Biom J 2006;48:286–301.

16 Peng EY, Lee MB, Tsai ST, Tsai ST, Yang CC, Morisky DE, Tsai LT, Weng YL, Lyu SY. Population-based post-crisis psychological distress: an example from the SARS outbreak in Taiwan. J Formos Med Assoc 2010;109:524–32.

17 Huckins JF, daSilva AW, Wang W, Hedlund E, Rogers C, Nepal SK, Wu J, Obuchi M, Murphy EI, Meyer ML, Dylan DW, Holtzheimer PE, Campbell AT. Mental health and behavior of college students during the early phases of the COVID-19 pandemic: longitudinal smartphone and ecological momentary assessment study. J Med Internet Res 2020;22:e20185.

18 Kutcher S, Wei Y. School mental health: a necessary component of youth mental health policy and plans. World Psychiatry 2020;19:174–5.

19 Cellini N, Canale N, Mioni G, Costa S. Changes in sleep pattern, sense of time and digital media use during COVID-19 lockdown in Italy. J Sleep Res 2020;e13074.

20 Koren D, Arnon I, Lavie P, Klein E. Sleep complaints as early predictors of posttraumatic stress disorder: a 1-year prospective study of injured survivors of motor vehicle accidents. Am J Psychiatry 2002;159:855–7.

21 Zhang J, Zhu S, Du C, Zhang Y. Posttraumatic stress disorder and somatic symptoms among child and adolescent survivors following the Lushan earthquake in China: A six-month longitudinal study. J Psychosom Res 2015;79:100–6.

